# Clinical and prognostic significance of the C-reactive protein-albumin-lymphocyte (CALLY) index in breast cancer: a systematic review and meta-analysis

**DOI:** 10.64898/2026.10.22.26366242

**Authors:** Kimia Jalalian Rad, Saeed Kadri, Abdullah Yakiti, Sevil Ghasemi, Hirbod hadizadeh moallem, Somayyeh Noei Teymoordash, Danyal Yarahmadi

**Author notes:** **Corresponding author**: Danyal Yarahmadi, Gastrointestinal and Liver Diseases Research Center, Iran University of Medical Sciences, Tehran, Iran.

## Abstract

**Background:** The C-reactive protein–albumin–lymphocyte (CALLY) index integrates inflammatory, nutritional and immune-related biomarkers and may offer an accessible measure of host status in breast cancer. However, its prognostic relevance and clinical value remain uncertain. This systematic review and meta-analysis evaluated its associations with survival and treatment response and examined predictive performance.

**Methods:** PubMed, Embase, Scopus and Web of Science were searched from inception to 1 October 2026, supplemented by reference-list and forward-citation searching. Studies evaluating CALLY in adults with invasive breast cancer were eligible. Multivariable-adjusted hazard ratios (HRs) comparing low versus high baseline CALLY were pooled using restricted maximum likelihood random-effects models with modified Hartung–Knapp intervals. Risk of bias and evidence certainty were assessed using the Newcastle–Ottawa Scale and GRADE, respectively.

**Results:** Nine publications included 2,568 reported patients. Four studies comprising 1,394 patients contributed to each survival synthesis. Low CALLY yielded pooled HRs of 2.03 for overall survival (95% CI 1.20-3.45; I²=72.4%) and 2.00 for disease-free survival (95% CI 0.91–4.41; I²=77.7%). Both intervals included no association. In one HER2-positive cohort, low baseline CALLY was associated with lower odds of pathological complete response (adjusted OR 0.25, 95% CI 0.12–0.50). Apparent overall-survival area under the receiver-operating-characteristic curve (AUC) ranged from 0.688 to 0.740. External validation and demonstrated clinical utility were absent.

**Conclusions:** CALLY shows potential prognostic and treatment-response associations, but imprecision, heterogeneity and limited validation prevent routine clinical adoption. Standardized measurement, prospective validation and demonstration of incremental clinical value are needed.

## Background

Breast cancer is the most frequently diagnosed malignancy and the leading cause of cancer-related mortality among women worldwide (1). Despite advances in surgery, chemotherapy, radiotherapy, targeted therapies, and endocrine treatment, breast cancer continues to be associated with considerable morbidity and mortality (2). In 2024, an estimated 2.4 million women were diagnosed with breast cancer, and approximately 694,000 deaths were attributed to the disease worldwide (3). Therefore, accessible and effective approaches are needed to improve risk stratification and predict survival outcomes in patients with breast cancer.

Although the Tumor–Node–Metastasis (TNM) staging system remains a cornerstone of prognostic assessment, anatomical staging alone may not adequately predict prognosis or guide individualized treatment, given that biological and molecular attributes of the tumor are not incorporated to this framework (4). The prognostic significance of nutritional and immune-inflammatory biomarkers has been increasingly recognized in cancer patients (5). In breast cancer several prognostic indices incorporating these parameters have been investigated, including the prognostic nutritional index, controlling nutritional status score, systemic immune-inflammation index, and pan-immune-inflammation value, all of which have been associated with survival outcomes (6–8). The C-reactive protein–albumin–lymphocyte (CALLY) index is a novel composite immuno-nutritional marker that combines indicators of systemic inflammation, nutritional status, and immune function into a single index (9). Its prognostic value has been investigated in several malignancies, including hepatocellular carcinoma, colorectal, gastric, nasopharyngeal, ovarian and breast cancer (9–14).

The CALLY index comprises three components. CRP is a well-established acute-phase inflammatory marker, and elevated serum CRP levels have been associated with tumor invasion, metastasis, and poor prognosis in breast cancer (15). Serum albumin reflects the nutritional status of cancer patients, while malnutrition may impair immune defense mechanisms and contribute to tumor progression (16). Lymphocytes are the main components of the antitumor immune response and play a protective role against tumors. Low lymphocyte counts are thought to reflect an inadequate immune response, which may increase the risk of tumor recurrence or metastasis (17, 18).

Although its prognostic value has been investigated in other malignancies, evidence in breast cancer remains limited and heterogeneous, and a comprehensive assessment of its association with breast cancer outcomes through pooled analysis is still lacking. Therefore, this systematic review and meta-analysis aims to assess the association between the CALLY index and overall survival (OS), disease-free survival (DFS), as well as its association with pathological complete response (pCR) following neoadjuvant therapy, in patients with breast cancer.

## Methods

### Study design and registration

This systematic review and meta-analysis evaluated the clinical and prognostic significance of the CALLY index in adults with breast cancer. Reporting followed the Preferred Reporting Items for Systematic Reviews and Meta-Analyses (PRISMA) 2020 statement (19). The protocol was registered in PROSPERO under registration number CRD20261533473. The PRISMA checklist is provided in Supplementary Table S1.

### Information sources and search strategy

PubMed/MEDLINE, Embase, Scopus and Web of Science Core Collection, were searched from inception to 1 October 2026. The search combined controlled vocabulary, where available, and free-text terms for breast malignancies with the CALLY acronym or its constituent biomarkers: C-reactive protein, albumin and lymphocytes. Search syntax was adapted to each database, and no language restrictions were applied. Complete search strategies, search dates and retrieval counts are provided in Supplementary Table S2. We also screened the reference lists of included studies and relevant reviews and conducted forward-citation searches of included studies to identify additional eligible reports.

### Eligibility criteria and study selection

Eligible studies evaluated the composite CALLY index in adults with established invasive breast cancer and reported at least one relevant clinical outcome, clinicopathological association or measure of predictive performance. Cohort and cross-sectional studies were eligible. The primary prognostic analysis concerned baseline CALLY and overall survival in patients with nonmetastatic disease; studies involving other disease settings remained eligible for the relevant secondary analyses. Studies evaluating pathological complete response following neoadjuvant systemic therapy, post-treatment CALLY or changes during treatment were also eligible. Investigations of individual biomarker components without evaluation of the composite index were excluded. Diagnostic accuracy for detecting breast cancer in screening or suspected-cancer populations was outside the review scope; discrimination between stages or nodal categories among patients with established breast cancer remained eligible. No minimum follow-up duration was required. Potentially overlapping reports were compared by institution, recruitment period and population characteristics, with duplicate patients contributing only once to a given synthesis.

Following deduplication using Endnote 21, titles and abstracts were screened, and potentially eligible reports underwent full-text assessment. Screening was performed by two reviewers independently, with disagreements resolved by third reviewer. Reasons for full-text exclusion were recorded.

### Outcomes and definitions

The primary outcome was OS and DFS. OS defined as time from the study-specified origin to death from any cause. DFS was extracted according to each study’s definition, including the time origin and event components, such as local or distant recurrence, second primary malignancy, and death, where specified. The primary effect measure was the multivariable-adjusted hazard ratio for low versus high baseline CALLY in nonmetastatic breast cancer.

Secondary outcomes included pathological complete response, clinicopathological associations, and endpoint-specific predictive performance. For pathological complete response, the reported definition was extracted, distinguishing breast-only response from breast-and-node response where possible. The preferred definition was ypT0/is ypN0. Baseline measurements were distinguished from post-treatment measurements and changes during therapy. Predictive performance included reported area under the receiver-operating-characteristic curve (AUC)s, time-dependent AUCs, C-indices, sensitivity and specificity. Calibration, internal or external validation, incremental value and decision-curve findings were extracted separately from measures of association.

### Data extraction and effect harmonization

Data were collected using a structured extraction form and extracted variables included publication details, study design, country, recruitment period, population size, disease stage, molecular subtype, treatment setting and demographic characteristics. CALLY-specific information included measurement timing, calculation formula, laboratory units, cutoff value, cutoff-selection method and category sizes.

For survival outcomes, extraction covered time origins, follow-up duration, event counts, hazard ratios, confidence intervals and adjustment variables. Multivariable-adjusted estimates were prioritized for the primary synthesis; unadjusted and continuous effects were distinguished from adjusted categorical comparisons. Categorical survival effects were harmonized to low versus high CALLY. Where a reported comparison was reversed, the hazard ratio and confidence limits were reciprocated, with the confidence-limit order reversed. Ambiguous reference categories were explicitly documented. CALLY-only estimates were distinguished from estimates for multicomponent scores and models containing CALLY.

For response outcomes, reported odds ratios and adjustment variables were extracted. Unadjusted odds ratios derived from available response/nonresponse counts were identified as calculated estimates. Conflicting numerical reports were retained with qualifications rather than treated as independent observations. Study-specific cutoffs were not assumed to be interchangeable across calculation units or clinical settings.

### Risk of bias and certainty of evidence

Methodological assessment used the Newcastle–Ottawa Scale (NOS) (20). The cohort version evaluated selection, comparability and outcome ascertainment, with a maximum score of nine. An adapted cross-sectional version, with a maximum score of ten, was used for cross-sectional analyses. Outcome-specific assessments were retained where analyses within a publication differed. Item-level judgments and scores are presented in Supplementary Table S5.

Certainty of evidence was assessed for OS, DFS and the association between baseline CALLY and pCR using GRADE guidance for prognostic-factor research (21). Evidence began at high certainty and was assessed for limitations concerning risk of bias, inconsistency, indirectness, imprecision and publication bias; observational design alone did not warrant downgrading. Outcome-specific ratings and supporting explanations are presented in Supplementary Table S6.

### Statistical analysis

Survival associations were synthesized on the natural logarithmic hazard-ratio scale. Standard errors were calculated from reported 95% confidence intervals as. Random-effects models used restricted maximum likelihood to estimate between-study variance. Pooled confidence intervals and hypothesis tests used the modified Hartung–Knapp approach, with the variance adjustment factor constrained to be at least one and a degrees of freedom. Estimates were back-transformed for presentation; hazard ratios above one indicated higher hazards in the low-CALLY group. OS was synthesized separately from recurrence-related outcomes. Between-study heterogeneity was summarized using, with between-study variance estimated by the random-effects model. Robustness was examined through leave-one-out analyses using the same model and confidence-interval method. With four studies per survival synthesis, funnel plots, formal small-study-effect tests and meta-regression were not undertaken. Response associations, dynamic-CALLY findings and predictive-performance measures were summarized descriptively because compatible estimates were insufficient for pooling. AUCs were not combined across different endpoints, prediction horizons, models or validation settings. Analyses were performed using R version 4.5.3 using the *meta* and *metafor*.

## Results

### Study selection

The database searches identified 542 records: 61 from PubMed, 201 from Embase, 187 from Scopus, and 93 from Web of Science. After removing 194 duplicates, 348 records underwent title and abstract screening, of which 323 were excluded. All 25 reports sought for retrieval were obtained and assessed for eligibility. Sixteen reports were excluded because of an ineligible population or outcome, absence of evaluation of the composite CALLY index, or an ineligible publication type or study design. Nine publications were included in the review, with four contributing to both the adjusted overall survival and DFS meta-analyses. Figure 1 presents the study-selection flow.

**Figure 1.**
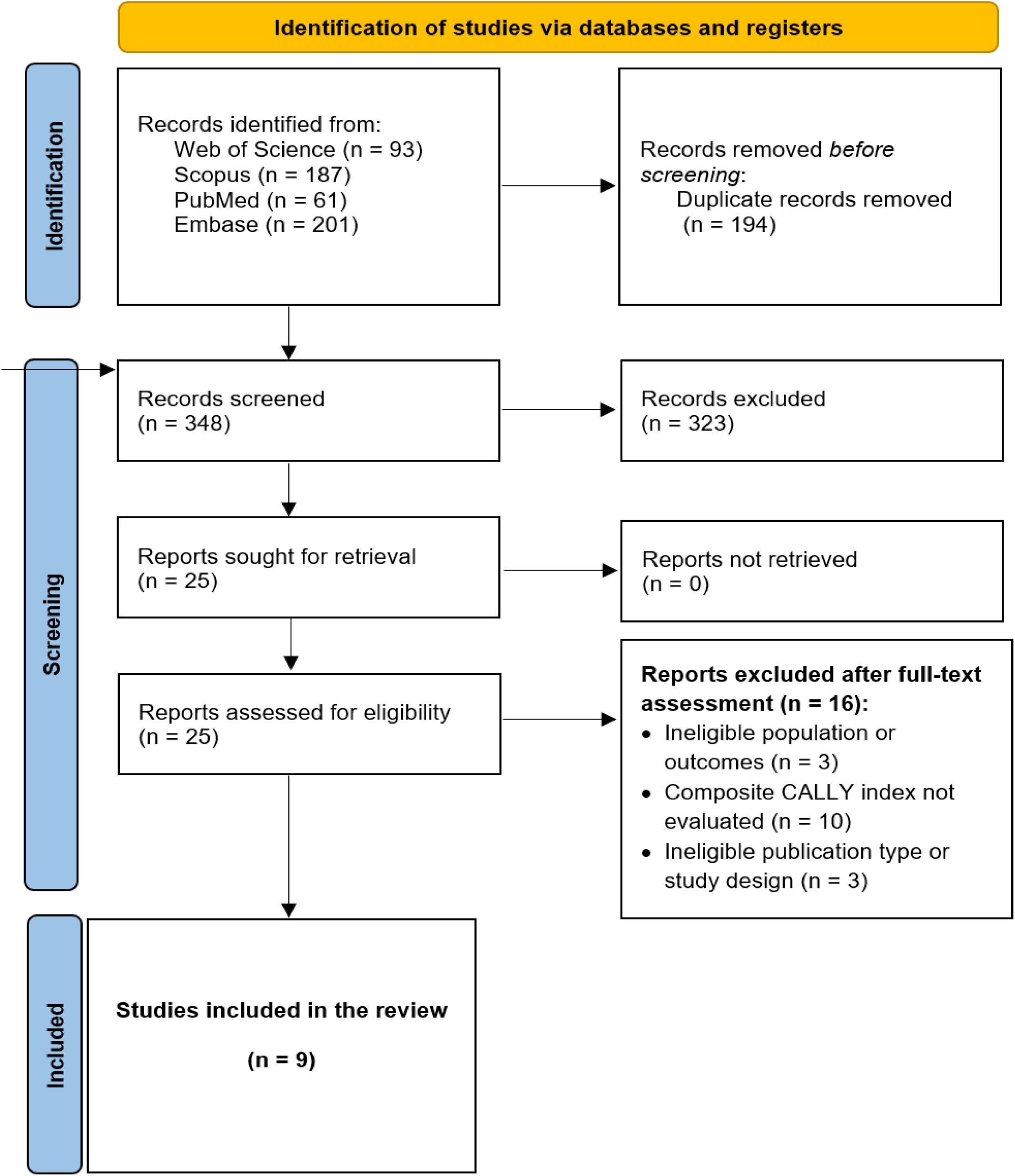
PRISMA 2020 flow diagram of the study selection process.

### Study characteristics and CALLY measurement

Nine publications contributed ten principal cancer cohorts or analysis populations, with a summed reported sample size of 2,568 patients(12, 22–28). Four reports were from Turkey, three from China, one from Iran, and one from Nigeria. Four retrospective surgical cohorts contributed adjusted survival estimates for 1,394 patients (12, 22, 24, 26). These included stage I–III disease, a stage III cohort and a triple-negative breast cancer cohort. One report evaluated baseline CALLY and neoadjuvant response, and one evaluated changes during neoadjuvant treatment; the remaining reports primarily examined clinicopathological or nodal associations. Recruitment, CALLY cutoffs, clinical populations and treatment characteristics are summarized in Table 1A–B.

**Table 1.** Study characteristics. Panel A: design, demographics and CALLY groups.

| Study | Country | Design | Recruitment period | Age, years: statistic (dispersion) | High / low CALLY, N | Cancer N |
| --- | --- | --- | --- | --- | --- | --- |
| Jimoh 2023 (27) | Nigeria | Hospital-based cross-sectional series | NR | NR | NA; continuous | 135 |
| Zhuang 2024 (12) | China | Retrospective surgical cohort | 2017–2018 | Median 50 (IQR 45–61; range 29–75) | 112 / 62 | 174 |
| Karatlı 2026 (29) | Turkey | Retrospective treatment-response cohort | 2023–2024 | Median 50.8 (range 25–79) | NR | 169 |
| Zhu 2025 (22) | China | Retrospective surgical cohort | 2010–2015 | Median 52 (range 26–79) | 133 / 54 | 187 |
| Algın 2026 (25) | Turkey | Cross-sectional stage association; retrospective records | 2018–2024 | Range 21–70; central value NR | 162 / 141 (derived) | 303 |
| Bahardoust 2026 (26) | Iran | Multicenter retrospective surgical cohort | 2011–2022 | Mean 48.2 ± 9.6 SD | 566 / 295 | 861 |
| Güneş 2026 (23) | Turkey | Retrospective cross-sectional nodal analyses | 2019–2026 | Upfront: mean 58.7 ± 11.4 SD; post-NACT: NR | NA; continuous | 247 + 80 |
| Güney 2026 (28) | Turkey | Retrospective consecutive treatment-response cohort | 2024 | Mean 52 ± 12 SD | Baseline NR; favorable/unfavorable dynamic: 132 / 108 | 240 |
| Liu 2026 (24) | China | Retrospective surgical cohort | 2011–2020 | Median 50 (range 28–80) | 89 / 83 | 172 |

NR, not reported; NA, not applicable; NACT, neoadjuvant systemic therapy; RT, radiotherapy; BCS, breast-conserving surgery; TNBC, triple-negative breast cancer; SLNB, sentinel lymph-node biopsy; ALND, axillary lymph-node dissection. Age retains each study’s reported statistic. Treatment counts may overlap. Güneş contributes two populations; Güney’s 132/108 groups describe CALLY change rather than baseline levels. Algn’s high/low counts were derived from reported complete stage separation. Zhuang’s chemotherapy subgroup total and Jimoh’s metastatic-status counts contain discrepancies. Cutoffs use study- specific calculation units and are not directly interchangeable.

**Table 1.**
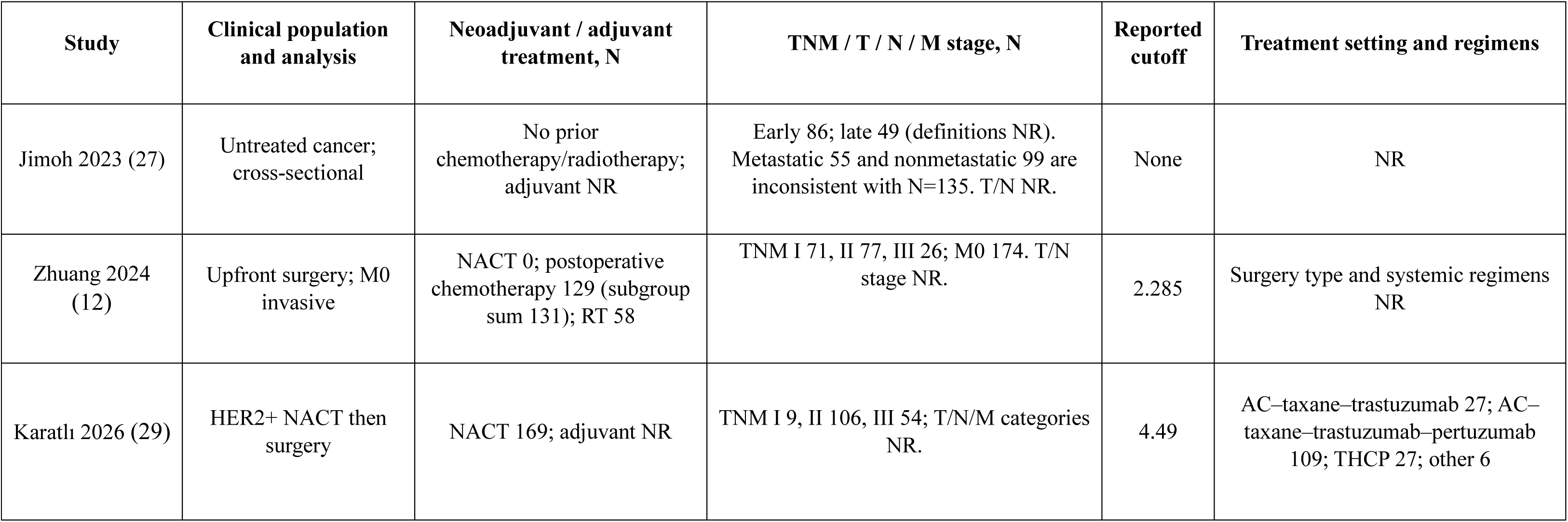
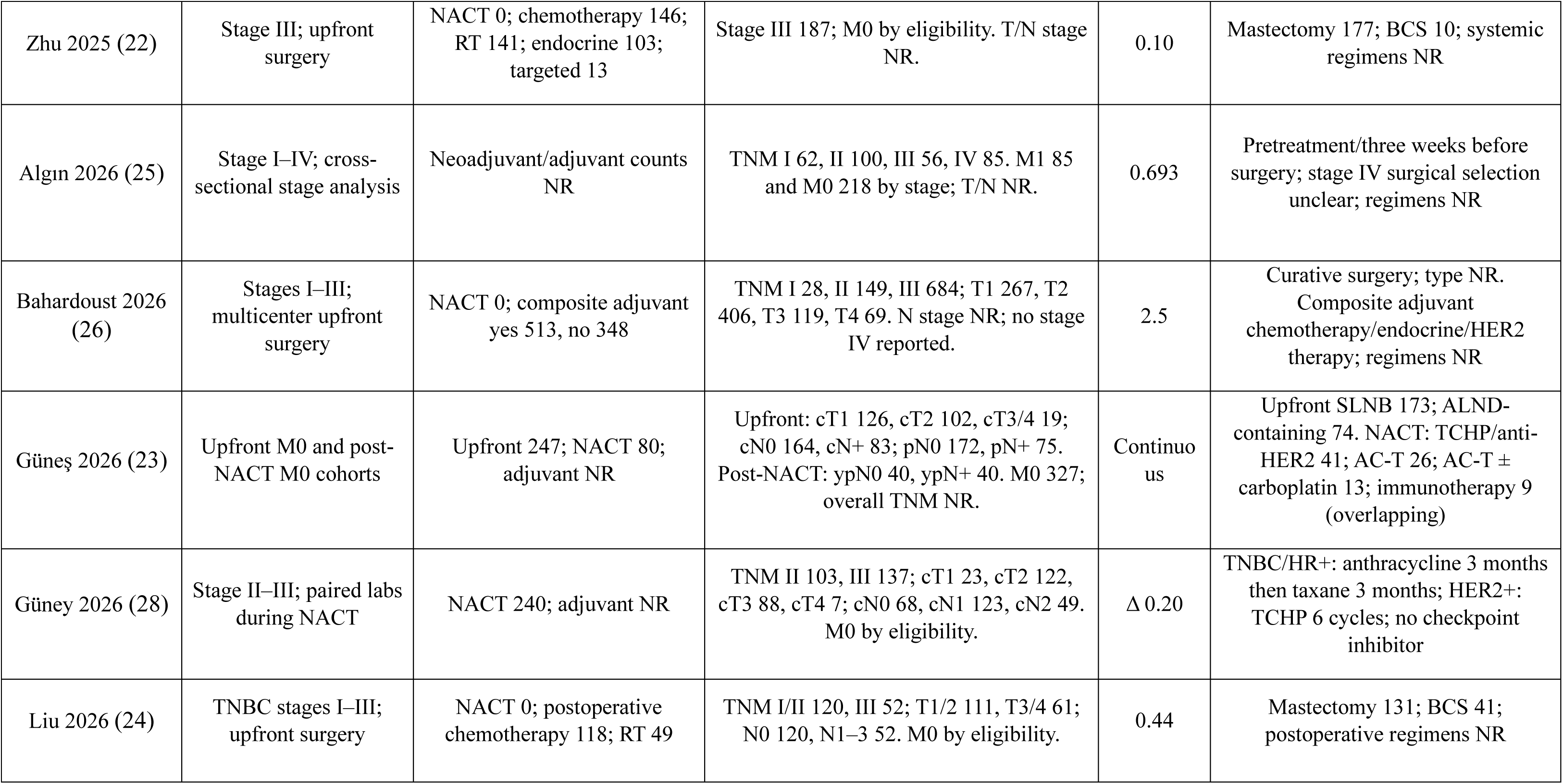
Panel B: clinical stage and treatment characteristics.

### Risk of bias

Seven outcome-specific cohort NOS assessments across six publications yielded scores of 5–7/9, with a median of 7. Five adapted cross-sectional assessments across three publications yielded scores of 4–8/10, with a median of 6 (Table S5). The cohort assessments consistently identified limitations in representativeness and completeness of follow-up, reflecting selection based on complete data or treatment completion and insufficient accounting for the source population and follow-up losses.

### Overall survival

Four studies contributed multivariable-adjusted OS estimates for baseline CALLY (12, 22, 24, 26). The pooled HR for low versus high CALLY was 2.03 (95% CI 1.20–3.45; P=0.079; 1,394 patients), with substantial heterogeneity (I²=72.4%; Figure 2).

**Figure 2.**
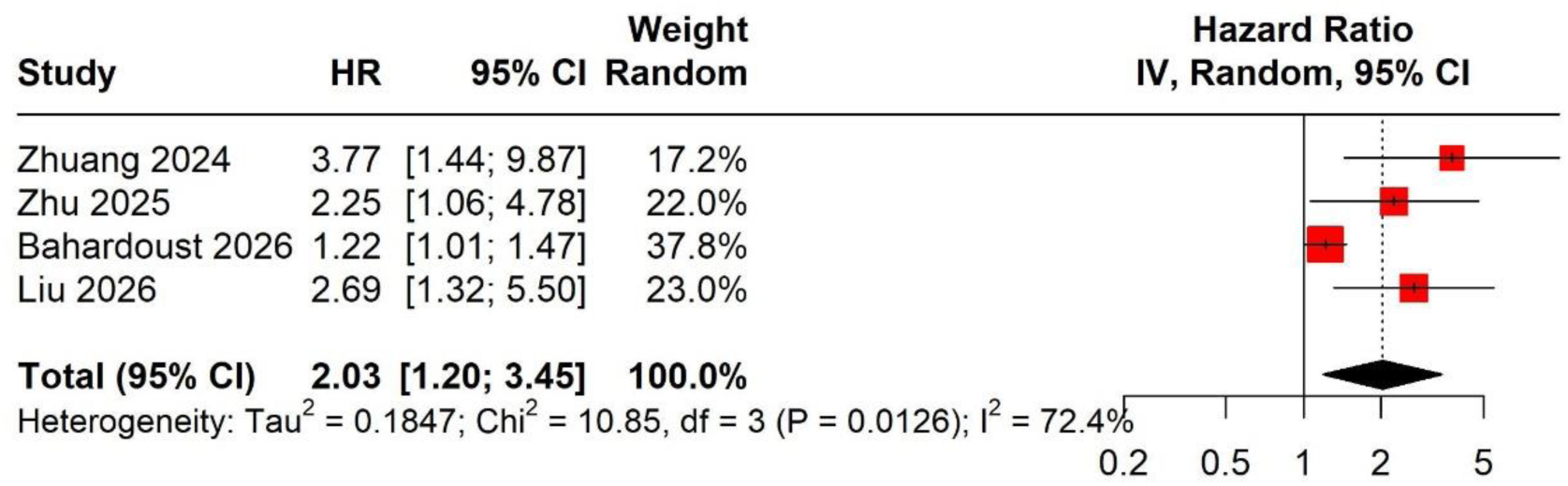
Adjusted overall survival meta-analysis.

### Disease-free and recurrence-free survival

Four studies reported adjusted DFS estimates (12, 22, 24, 26). The DFS analysis included 1,394 patients and yielded an HR of 2.01 for low versus high baseline CALLY (95% CI 1.23–3.26; P=0.037; I²=77.7%; Figure 3).

**Figure 3.**
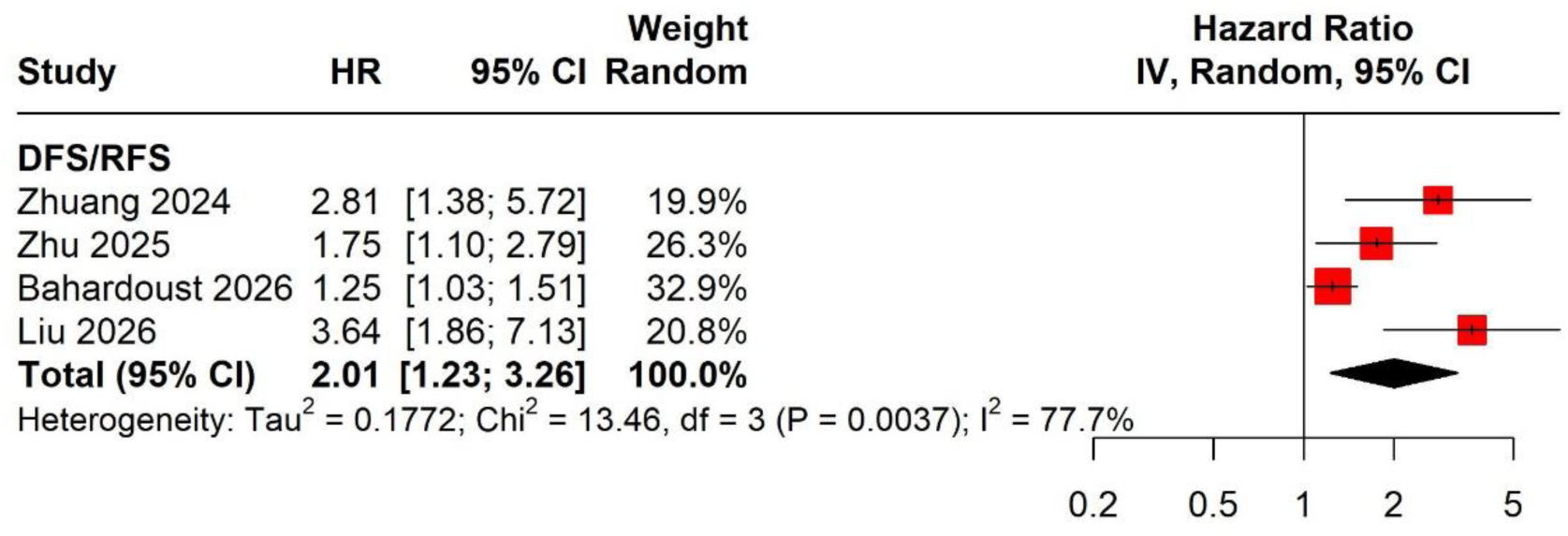
Adjusted DFS meta-analysis.

### Pathological complete response and dynamic CALLY

Karatlı et al. included 169 patients with HER2-positive breast cancer, of whom 96 achieved pCR (29). High versus low baseline CALLY was associated with pCR after adjustment for ER status, Ki-67 and regimen (OR 4.03, 95% CI 1.99–8.18), equivalent to an OR of 0.25 (95% CI 0.12–0.50) for low versus high CALLY (Table S3). The exact breast and nodal components of the pCR definition were not reported. This was the only adjusted baseline CALLY–pCR association study, so no corresponding meta-analysis was performed.

Güney et al. evaluated 240 neoadjuvant-treated patients, including 94 with ypT0/Tis ypN0 pCR (28). Favorable versus unfavorable CALLY change groups had pCR in 75/132 and 19/108 patients, respectively, yielding a derived unadjusted OR of 6.16 (95% CI 3.37–11.27). A model combining CALLY change with NPAR change reported an adjusted OR of 23.5 (95% CI 11.4–48.6). Its comparison was unspecified, and the combined-score counts conflicted with the component marginals. Dynamic CALLY and combined-score associations were kept separate from baseline CALLY findings (Table S3).

### Discrimination calibration and incremental value

CALLY-only discrimination varied by endpoint and setting (Table 2; Table S4). Apparent OS AUCs were 0.730 (95% CI 0.637–0.824), 0.740 (0.660–0.810) and 0.688 (0.603–0.773) in Zhuang, Bahardoust and Liu, respectively (12, 24, 26). Their time horizons and handling of censoring were insufficiently described for AUC pooling. Zhu reported an AUC of 0.604 (0.429–0.809) without an explicit ROC target (22). Baseline pCR AUC was 0.681 (0.597–0.765), while change-based pCR AUC was 0.685 without a confidence interval (28). Upfront and post-neoadjuvant nodal AUCs were 0.576 and 0.593 (23). Algın reported an apparent AUC of 1.00 for stage III–IV versus stage I–II discrimination, without independent validation of the threshold (25).

**Table 2.**
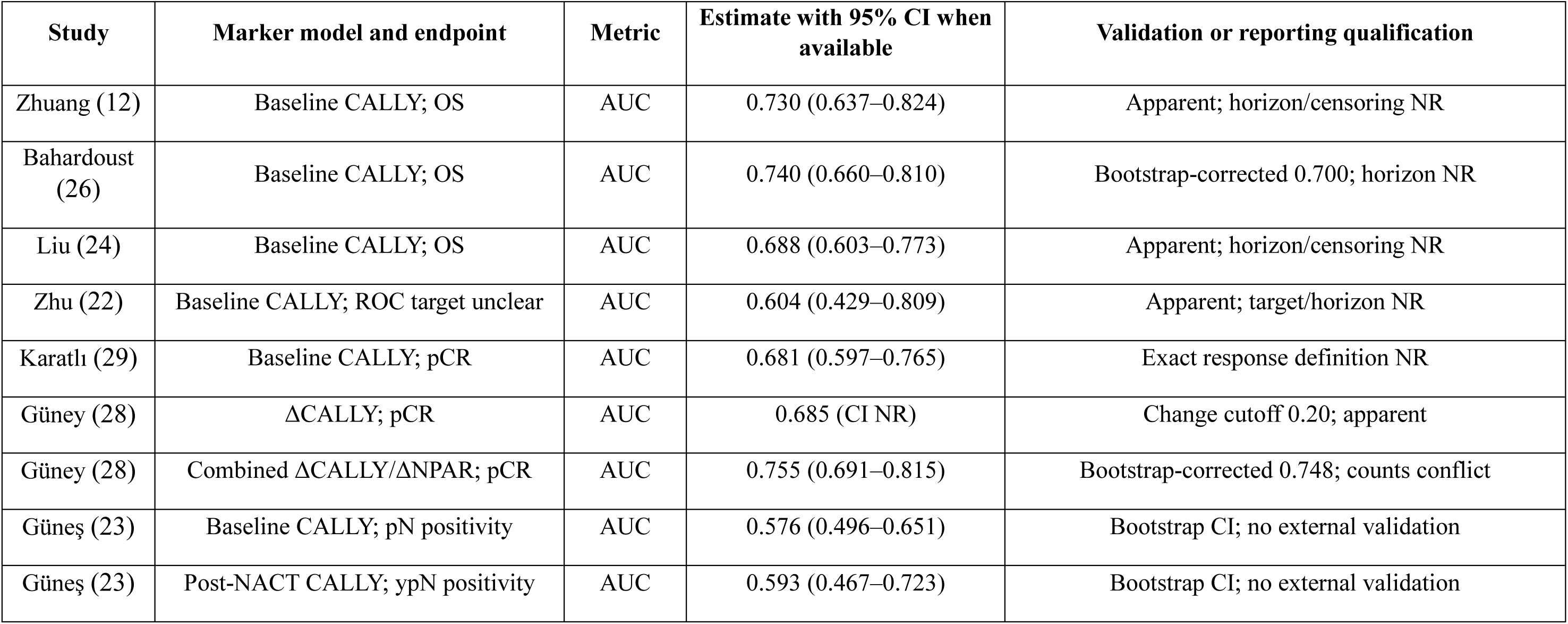
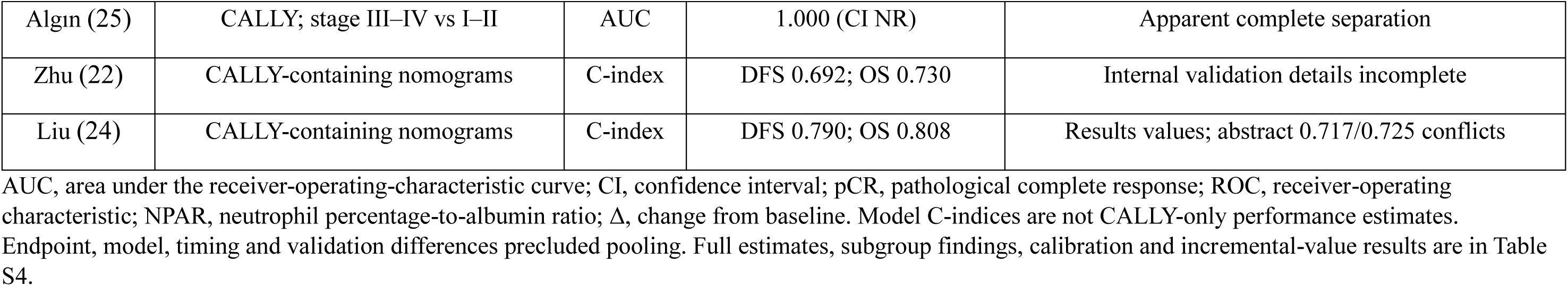
Discriminative performance by endpoint and model.

CALLY-containing nomograms in Zhu had C-indices of 0.692 for DFS and 0.730 for OS (18). Liu reported Results-section C-indices of 0.790 and 0.808, but its abstract gave 0.717 and 0.725; these were retained as conflicting reports (24). Both studies presented graphical calibration and decision curves, without sufficient numerical calibration or incremental-value information for synthesis. Güney’s combined response score had an apparent AUC of 0.755 and bootstrap-corrected AUC of 0.748; these applied to the combined model, not CALLY alone (28). Adding CALLY to Güneş’s clinical model changed AUC by −0.0001 (95% CI −0.0013 to 0.0041), while the Brier score changed from 0.0271 to 0.0260 (23). No external validation or clinical-impact study demonstrating improved outcomes from CALLY-guided decisions was identified.

### Sensitivity analyses and reporting bias

Leave-one-out analyses remained imprecise: OS point estimates ranged from 1.76 to 2.72, and DFS estimates from 1.65 to 2.47; all modified Hartung–Knapp confidence intervals included 1. With four studies per adjusted survival analysis, funnel plots, formal small-study-effect tests and meta-regression were not undertaken. Publication bias could not be reliably excluded.

### Certainty of evidence

Certainty of evidence was low for OS and very low for the exploratory DFS analysis. Both outcomes were limited by risk of bias and imprecision, while differences in recurrence-related endpoint definitions additionally reduced certainty for DFS. Certainty was low for the association between baseline CALLY and pCR in HER2-positive breast cancer because of risk of bias and incomplete specification of the response definition. Outcome-specific judgments and reasons for downgrading are presented in Table S6.

## Discussion

This systematic review and meta-analysis found that lower baseline CALLY values tended to be associated with poorer breast cancer outcomes. However, the evidence remains too limited and inconsistent to support routine clinical use. Nine publications examined CALLY in relation to survival, treatment response, and clinicopathological characteristics. Low CALLY was associated with approximately twice the risk of death and DFS.

The survival studies generally showed associations in the same direction, but their populations differed considerably. They included patients with nonmetastatic disease, stage III disease, and triple-negative breast cancer (12, 22, 24, 26). Differences in disease stage, tumor biology, treatment and statistical adjustment may explain the substantial heterogeneity observed for overall survival (I²=72.4%). The recurrence analysis was similarly heterogeneous (I²=77.7%) and disease-free and recurrence-free survival definitions that were not fully comparable. Its pooled estimate should therefore be interpreted as an exploratory summary of related recurrence outcomes rather than a standardized disease-free survival effect.

The possible relationship between CALLY and breast cancer outcomes has a reasonable biological explanation. CALLY combines CRP, albumin, and lymphocyte count, which together provide information about inflammation, the patient’s general condition, and immune function. Previous studies have linked CALLY to survival in hepatocellular carcinoma and elevated CRP to poorer survival in breast cancer (9, 30). However, CALLY is not specific to cancer. Albumin levels can fall because of inflammation and do not directly indicate malnutrition (31), while blood lymphocyte counts may not reflect immune activity within the tumor (32). Therefore, low CALLY may be a general sign of inflammation, greater disease burden, or other health problems rather than a direct cause of poorer outcomes.

The evidence linking CALLY to response to neoadjuvant treatment is encouraging but remains limited. In a study of 169 patients with HER2-positive breast cancer, higher pretreatment CALLY was independently associated with a greater likelihood of achieving pathological complete response. However, its predictive ability was modest (AUC 0.681), and the definition of pCR was not clearly reported (29). Furthermore, achieving pCR does not automatically confirm a survival benefit or show which treatment is most suitable for an individual patient (33). Another study reported that favourable changes in CALLY during treatment were associated with pCR (28), but the CALLY-only analysis was unadjusted, while the adjusted result related to a combined score. Changes measured during or after treatment may also be influenced by treatment effects, toxicity, infection, or supportive care. Therefore, dynamic CALLY should be assessed separately from pretreatment CALLY.

Findings for clinicopathological features varied between studies. Lower CALLY values were reported in patients with more advanced or unfavorable disease characteristics (25, 27), but these cross-sectional results cannot confirm that CALLY predicts future outcomes. Algın et al. reported perfect discrimination between early and advanced disease (AUC 1.00), which is unusual and requires confirmation in an independent population (25). In comparison, Güneş et al. found that CALLY had limited ability to identify nodal involvement and added little to established clinical predictors (23). Overall, the usefulness of CALLY appears to vary according to the patient population and outcome being studied, so findings from one clinical setting should not be applied directly to another.

A statistical association does not necessarily mean that CALLY can accurately predict outcomes for individual patients. CALLY alone showed only modest discrimination for overall survival, with AUC values ranging from approximately 0.69 to 0.74 (12, 24, 26), and these results could not be pooled because prediction periods and the handling of censored data were not consistently reported. Although some models containing CALLY showed favorable discrimination, calibration, and decision-curve results (22, 24), they also included established clinical and pathological factors, making it difficult to determine the additional value of CALLY itself. Clinical use is further limited by the wide variation in reported cutoffs, which ranged from 0.10 to 4.49 and differed according to calculation methods, laboratory units, patient populations, and measurement timing (12, 22, 24–26, 29). These thresholds cannot be directly compared or combined into a universal cutoff. Future studies should clearly report how and when CALLY is calculated, analyze it initially as a continuous measure, assess whether it adds value beyond established predictors, and validate any proposed cutoff in an independent population, in accordance with REMARK recommendations (34).

A key strength of this review was its specific focus on breast cancer and its separate assessment of survival, baseline treatment response, changes in CALLY during treatment, and predictive performance. However, most included studies were retrospective, and the survival evidence came from a limited range of countries, reducing the generalizability of the findings. Differences in patient selection, follow-up reporting and statistical adjustment also made comparisons between studies difficult and left the results vulnerable to residual confounding. Finally, the small number of available studies prevented a reliable assessment of publication bias, meaning that studies with favorable results may be overrepresented.

Future research should include prospective, multicenter studies using a consistent method for calculating CALLY and clearly defined timing of blood collection before treatment. Studies should identify important confounders in advance, report outcome events, manage missing data appropriately and assess both discrimination and calibration at clinically relevant time points. CALLY should also be compared directly with established predictors and with its individual components, followed by validation in independent populations (34, 35). Research examining changes during treatment should use standard measurement intervals and account for treatment effects, toxicity, and infection. Finally, clinical-impact studies are needed to determine whether adding CALLY to current assessments improves clinical decisions or patient outcomes.

## Conclusion

CALLY is a simple and potentially useful marker for identifying patients with breast cancer who may have poorer outcomes. However, the current evidence is limited by imprecise estimates, substantial differences between studies, and insufficient evidence that CALLY improves existing prediction methods. Therefore, CALLY is not yet ready for routine clinical use and requires standardised measurement, independent validation and evidence of clear clinical benefit.

## Abbreviations

CALLY: C-reactive protein-albumin-lymphocyte
HR: Hazard ratio
CI: Confidence interval
OR: Odds ratio
AUC: Area under the receiver-operating-characteristic curve
TNM: Tumor-Node-Metastasis
OS: Overall survival
DFS: Disease free survival
RFS: Recurrence free survival
pCR: Pathologic complete response
PRISMA: Preferred Reporting Items for Systematic Reviews and Meta-Analyses
NOS: Newcastle–Ottawa Scale
GRADE: Grading of Recommendations Assessment, Development and Evaluation
REMARK: Reporting Recommendations for Tumor Marker Prognostic Studies
REML: Residual maximum likelihood
ER: Estrogen receptor
HER2: Human epidermal growth factor receptor 2
NPAR: Neutrophil percentage to albumin ratio

## Acknowledgements

NA.

## Funding

Not applicable.

## Data availability

The datasets generated or analyzed during the current study are available from the corresponding author upon reasonable request.

## Author contributions (CRediT)

M.S.: Conceptualization, Methodology, Formal analysis, Writing – original draft, Writing – review & editing. S.K.: Methodology, Validation, Writing – review & editing. K.J.R.: Investigation, Data curation, Writing – original draft. A.Y.: Investigation, Data curation, Writing – original draft. S.G.: Investigation, Data curation, Writing – review & editing. H.H.M.: Investigation, Data curation, Writing – review & editing. S.N.T.: Validation, Supervision, Writing – review & editing. D.Y.: Conceptualization, Methodology, Supervision, Writing – review & editing. All authors reviewed and approved the final manuscript and agreed to be accountable for all aspects of the work.

## Declarations

### Human Ethics and Consent to Participate declarations

Not applicable.

### Ethics approval and consent to participate

Not applicable. This study is a systematic review and meta-analysis of published, de-identified data and did not involve direct contact with human participants.

## Consent to participate

Not applicable.

## Consent for publication

Not applicable.

## Protocol registration

PROSPERO CRD20261533473.

## Conflict of Interest Statement

The authors declare no conflicts of interest related to this work.

## Notes

### Competing Interest Statement

The authors have declared no competing interest.

