## Supplementary Material for "Clinical and prognostic significance of the C-reactive protein-albumin-lymphocyte (CALLY) index in breast cancer: a systematic review and meta-analysis"

**Table S1: PRISMA Checklist.**

| **Section and Topic** | **Item #** | **Checklist item** | **Location where item is reported** |
| --- | --- | --- | --- |
| **TITLE** | | |  |
| Title | 1 | Identify the report as a systematic review. | P1 |
| **ABSTRACT** | | |  |
| Abstract | 2 | See the PRISMA 2020 for Abstracts checklist. | P1 |
| **INTRODUCTION** | | |  |
| Rationale | 3 | Describe the rationale for the review in the context of existing knowledge. | P2 |
| Objectives | 4 | Provide an explicit statement of the objective(s) or question(s) the review addresses. | P3 |
| **METHODS** | | |  |
| Eligibility criteria | 5 | Specify the inclusion and exclusion criteria for the review and how studies were grouped for the syntheses. | P3 |
| Information sources | 6 | Specify all databases, registers, websites, organisations, reference lists and other sources searched or consulted to identify studies. Specify the date when each source was last searched or consulted. | P3 |
| Search strategy | 7 | Present the full search strategies for all databases, registers and websites, including any filters and limits used. | P3 |
| Selection process | 8 | Specify the methods used to decide whether a study met the inclusion criteria of the review, including how many reviewers screened each record and each report retrieved, whether they worked independently, and if applicable, details of automation tools used in the process. | P4 |
| Data collection process | 9 | Specify the methods used to collect data from reports, including how many reviewers collected data from each report, whether they worked independently, any processes for obtaining or confirming data from study investigators, and if applicable, details of automation tools used in the process. | P4 |
| Data items | 10a | List and define all outcomes for which data were sought. Specify whether all results that were compatible with each outcome domain in each study were sought (e.g. for all measures, time points, analyses), and if not, the methods used to decide which results to collect. | P4 |
|  | 10b | List and define all other variables for which data were sought (e.g. participant and intervention characteristics, funding sources). Describe any assumptions made about any missing or unclear information. | PP 4,5 |
| Study risk of bias assessment | 11 | Specify the methods used to assess risk of bias in the included studies, including details of the tool(s) used, how many reviewers assessed each study and whether they worked independently, and if applicable, details of automation tools used in the process. | P5 |
| Effect measures | 12 | Specify for each outcome the effect measure(s) (e.g. risk ratio, mean difference) used in the synthesis or presentation of results. | P5 |
| Synthesis methods | 13a | Describe the processes used to decide which studies were eligible for each synthesis (e.g. tabulating the study intervention characteristics and comparing against the planned groups for each synthesis (item #5)). | P5 |
|  | 13b | Describe any methods required to prepare the data for presentation or synthesis, such as handling of missing summary statistics, or data conversions. | P5 |
|  | 13c | Describe any methods used to tabulate or visually display results of individual studies and syntheses. | P5 |
|  | 13d | Describe any methods used to synthesize results and provide a rationale for the choice(s). If meta-analysis was performed, describe the model(s), method(s) to identify the presence and extent of statistical heterogeneity, and software package(s) used. | P5 |
|  | 13e | Describe any methods used to explore possible causes of heterogeneity among study results (e.g. subgroup analysis, meta-regression). | P5 |
|  | 13f | Describe any sensitivity analyses conducted to assess robustness of the synthesized results. | P5 |
| Reporting bias assessment | 14 | Describe any methods used to assess risk of bias due to missing results in a synthesis (arising from reporting biases). | P5 |
| Certainty assessment | 15 | Describe any methods used to assess certainty (or confidence) in the body of evidence for an outcome. | P5 |
| **RESULTS** | | |  |
| Study selection | 16a | Describe the results of the search and selection process, from the number of records identified in the search to the number of studies included in the review, ideally using a flow diagram. | P6 |
|  | 16b | Cite studies that might appear to meet the inclusion criteria, but which were excluded, and explain why they were excluded. | P6 |
| Study characteristics | 17 | Cite each included study and present its characteristics. | P6 |
| Risk of bias in studies | 18 | Present assessments of risk of bias for each included study. | P6 |
| Results of individual studies | 19 | For all outcomes, present, for each study: (a) summary statistics for each group (where appropriate) and (b) an effect estimate and its precision (e.g. confidence/credible interval), ideally using structured tables or plots. | P7 |
| Results of syntheses | 20a | For each synthesis, briefly summarise the characteristics and risk of bias among contributing studies. | Table S5 |
|  | 20b | Present results of all statistical syntheses conducted. If meta-analysis was done, present for each the summary estimate and its precision (e.g. confidence/credible interval) and measures of statistical heterogeneity. If comparing groups, describe the direction of the effect. | P7 |
|  | 20c | Present results of all investigations of possible causes of heterogeneity among study results. | PP 8,9 |
|  | 20d | Present results of all sensitivity analyses conducted to assess the robustness of the synthesized results. | P8 |
| Reporting biases | 21 | Present assessments of risk of bias due to missing results (arising from reporting biases) for each synthesis assessed. | P8 |
| Certainty of evidence | 22 | Present assessments of certainty (or confidence) in the body of evidence for each outcome assessed. | PP 7,8 |
| **DISCUSSION** | | |  |
| Discussion | 23a | Provide a general interpretation of the results in the context of other evidence. | P8 |
|  | 23b | Discuss any limitations of the evidence included in the review. | P10 |
|  | 23c | Discuss any limitations of the review processes used. | P10 |
|  | 23d | Discuss implications of the results for practice, policy, and future research. | P10 |
| **OTHER INFORMATION** | | |  |
| Registration and protocol | 24a | Provide registration information for the review, including register name and registration number, or state that the review was not registered. | P3 |
|  | 24b | Indicate where the review protocol can be accessed, or state that a protocol was not prepared. | P19 |
|  | 24c | Describe and explain any amendments to information provided at registration or in the protocol. | NA |
| Support | 25 | Describe sources of financial or non-financial support for the review, and the role of the funders or sponsors in the review. | NA |
| Competing interests | 26 | Declare any competing interests of review authors. | P12 |
| Availability of data, code and other materials | 27 | Report which of the following are publicly available and where they can be found: template data collection forms; data extracted from included studies; data used for all analyses; analytic code; any other materials used in the review. | P11 |

Table S2. Search Strategy

| Database | Search Strategy |
| --- | --- |
| PubMed | ("Breast Neoplasms"[Mesh] OR ((breast[tiab] OR mammary[tiab]) AND (cancer*[tiab] OR neoplasm*[tiab] OR carcinoma*[tiab] OR malignan*[tiab] OR tumor*[tiab] OR tumour*[tiab]))) AND (CALLY[tiab] OR (("C-Reactive Protein"[Mesh] OR "C reactive protein"[tiab] OR "C-reactive protein"[tiab] OR CRP[tiab]) AND ("Albumins"[Mesh] OR albumin*[tiab]) AND ("Lymphocytes"[Mesh] OR lymphocyt*[tiab]))) |
| Embase | ('breast cancer'/exp OR ((breast OR mammary) AND (cancer* OR neoplasm* OR carcinoma* OR malignan* OR tumor* OR tumour*)):ti,ab,kw) AND (cally:ti,ab,kw OR (('c reactive protein'/exp OR 'c reactive protein':ti,ab,kw OR 'c-reactive protein':ti,ab,kw OR crp:ti,ab,kw) AND ('albumin'/exp OR albumin*:ti,ab,kw) AND ('lymphocyte'/exp OR lymphocyt*:ti,ab,kw))) |
| Scopus | TITLE-ABS-KEY((breast OR mammary) AND (cancer* OR neoplasm* OR carcinoma* OR malignan* OR tumor* OR tumour*)) AND TITLE-ABS-KEY(CALLY OR (("C reactive protein" OR "C-reactive protein" OR CRP) AND albumin* AND lymphocyt*)) |
| Web of Science | TS=((breast OR mammary) AND (cancer* OR neoplasm* OR carcinoma* OR malignan* OR tumor* OR tumour*)) AND TS=(CALLY OR (("C reactive protein" OR "C-reactive protein" OR CRP) AND albumin* AND lymphocyt*)) |

**Table S3 Treatment response, dynamic CALLY and other non-survival associations**

| **Study cohort and endpoint** | **Timing and reported comparison** | **Reported effect with 95% CI** | **Model and covariates** | **Analysis N and events** |
| --- | --- | --- | --- | --- |
| **Karatlı 2025 (1); HER2+ NACT; pCR** | Pretreatment; High vs low | OR 4.28 (2.24–8.19) | Unadjusted; None | 169; events 96.0 |
| **Karatlı 2025 (1); HER2+ NACT; pCR** | Pretreatment; High vs low | OR 4.03 (1.99–8.18) | Adjusted; CALLY; Ki67; ER; regimen | 169; events 96.0 |
| **Güneş 2026 (2); Upfront M0; pN positivity** | Baseline; Per CALLY doubling | OR per doubling 0.87 (0.71–1.06) | Unadjusted; None | 247; events 75.0 |
| **Güneş 2026 (2); Upfront M0; pN positivity** | Baseline; Per CALLY doubling | OR per doubling 0.89 (0.72–1.10) | Adjusted; Age per10y; cT | 247; events 75.0 |
| **Güneş 2026 (2); Upfront M0; pN positivity** | Baseline; Per CALLY doubling | OR per doubling 0.78 (0.51–1.21) | Adjusted Firth; Age per10y; cT; cN | 247; events 75.0 |
| **Güneş 2026 (2); Upfront M0; pN positivity** | Baseline; Per CALLY doubling | OR per doubling 0.89 (0.72–1.09) | Adjusted sensitivity; Age; cT; BMI | 247; events 75.0 |
| **Güneş 2026 (2); Upfront M0; pN positivity** | Baseline; Per CALLY doubling | OR per doubling 0.86 (0.69–1.08) | Adjusted sensitivity; Age; cT; BMI; smoking; cardiometabolic count; inflammation modifier | 247; events 75.0 |
| **Güneş 2026 (2); Upfront M0 inflammation restricted; pN positivity** | Baseline; Per CALLY doubling | OR per doubling 0.91 (0.73–1.15) | Adjusted sensitivity; Age; cT; BMI; smoking; cardiometabolic count; restriction | 224; events None |
| **Güneş 2026 (2); Neoadjuvant M0; Residual ypN positivity** | Post-treatment; Per CALLY doubling | OR per doubling 0.86 (0.68–1.09) | Unadjusted; None | 80; events 40.0 |
| **Güneş 2026 (2); Upfront node-positive; Observed >=4 positive nodes** | Baseline; Per CALLY doubling | OR per doubling 1.19 (0.74–1.92) | Unadjusted; None | 75; events 7.0 |
| **Güneş 2026 (2); Neoadjuvant node-positive; Observed >=4 positive nodes** | Post-treatment; Per CALLY doubling | OR per doubling 1.14 (0.80–1.64) | Unadjusted; None | 40; events 11.0 |
| **Güneş 2026 (2); Upfront M0; ALND-containing procedure** | Baseline; Per CALLY doubling | OR per doubling 0.97 (0.79–1.20) | Adjusted sensitivity; cT | 247; events 74.0 |
| **Güneş 2026 (2); Upfront node-positive ALND; Observed >=4 positive nodes** | Baseline; Per CALLY doubling | OR per doubling 1.08 (0.66–1.79) | Unadjusted sensitivity; None | 45; events 7.0 |
| **Güney 2026 (3); Paired NACT; pCR** | Delta post-pre; Favorable vs unfavorable deltaCALLY | OR 6.16 (3.37–11.27) | Unadjusted; None | 240; events 94.0 |
| **Güney 2026 (3); Paired NACT; pCR** | Changes in CALLY AND NPAR; Combined score contrast NR | OR 23.50 (11.40–48.60) | Adjusted; Subtype; cT3-4; cNpositive; combined score | 240; events 94.0 |

Karatlı’s adjusted pCR OR oriented low versus high is 0.248 (95% CI 0.122–0.503). Güney’s CALLY-change-only OR is derived from 75/57 versus 19/89 pCR/non-pCR counts; its adjusted combined-score result is not an adjusted single-CALLY effect. Nodal positivity and observed nodal burden are distinct endpoints. Per-doubling effects are not categorical low/high contrasts. No compatible multi-study response or dynamic-CALLY pool was available.

**Table S4. Endpoint-specific performance, calibration and incremental value**

| **Study and setting** | **Endpoint and model** | **Metric** | **Estimate with 95% CI if available** | **Validation** |
| --- | --- | --- | --- | --- |
| **Karatlı 2025 (1); HER2+ NACT** | pCR; CALLY | AUC | 0.681 (0.597–0.765) | Apparent; no validation reported |
| **Algın 2026 (4); Stages I-IV** | Advanced III-IV vs early I-II; CALLY | AUC | 1.000 (CI NR) | Apparent cutoff; no independent cohort |
| **Algın 2026 (4); MLP training** | 4-category stage; CRP, CALLY, albumin, lymphocytes, CA15-3 | Accuracy | 0.802 (CI NR) | 70% training; N217 derived from matrix |
| **Algın 2026 (4); MLP test** | 4-category stage; Same composite predictors | Accuracy | 0.849 (CI NR) | 30% internal holdout; N86 derived from matrix |
| **Güneş 2026 (2); Upfront M0** | pN positivity; CALLY | AUC | 0.576 (0.496–0.651) | 2000 bootstrap CI; apparent |
| **Güneş 2026 (2); post-NACT M0** | Residual ypN positivity; CALLY | AUC | 0.593 (0.467–0.723) | 2000 bootstrap CI; apparent |
| **Güneş 2026 (2); Upfront node-positive** | Observed >=4 nodes; CALLY | AUC | 0.433 (0.214–0.685) | 2000 bootstrap CI; apparent |
| **Güneş 2026 (2); post-NACT node-positive** | Observed >=4 nodes; CALLY | AUC | 0.476 (0.265–0.690) | 2000 bootstrap CI; apparent |
| **Güneş 2026 (2); Upfront node-positive ALND** | Observed >=4 nodes; CALLY | AUC | 0.451 (0.222–0.705) | 2000 bootstrap CI; apparent |
| **Güneş 2026 (2); Upfront M0** | pN positivity; Age+cT | AUC | 0.670 (CI NR) | 3000 paired bootstrap delta CI; no external validation |
| **Güneş 2026 (2); Upfront M0** | pN positivity; Age+cT+CALLY | AUC | 0.698 (CI NR) | 3000 paired bootstrap delta CI; no external validation |
| **Güneş 2026 (2); Upfront M0** | pN positivity; Add CALLY to age+cT | Delta AUC | 0.0270 (-0.0070–0.0690) | 3000 paired bootstrap delta CI; no external validation |
| **Güneş 2026 (2); Upfront M0** | pN positivity; Age+cT+cN | AUC | 0.988 (0.979–0.998) | 3000 paired bootstrap delta CI; no external validation |
| **Güneş 2026 (2); Upfront M0** | pN positivity; Age+cT+cN+CALLY | AUC | 0.988 (0.980–0.999) | 3000 paired bootstrap delta CI; no external validation |
| **Güneş 2026 (2); Upfront M0** | pN positivity; Add CALLY to age+cT+cN | Delta AUC | -0.0001 (-0.0013–0.0041) | 3000 paired bootstrap delta CI; no external validation |
| **Güneş 2026 (2); Upfront M0** | pN positivity; Age+cT+cN | Brier score | 0.0271 (CI NR) | 3000 paired bootstrap delta CI; no external validation |
| **Güneş 2026 (2); Upfront M0** | pN positivity; Age+cT+cN+CALLY | Brier score | 0.0260 (CI NR) | 3000 paired bootstrap delta CI; no external validation |
| **Bahardoust 2026 (5); All** | OS; horizon NR; CALLY | AUC | 0.740 (0.660–0.810) | Apparent; cutoff selected on same dataset |
| **Bahardoust 2026 (5); All** | OS; horizon NR; CALLY | Bias-corrected AUC | 0.700 (CI NR) | Bootstrap; resampling detail incomplete |
| **Bahardoust 2026 (5); All** | Survival model calibration; Multivariable model | Gronnesby-Borgan P | 0.950 (CI NR) | Internal assessment |
| **Zhuang 2024 (6); Upfront M0** | OS; horizon NR; CALLY | AUC | 0.730 (0.637–0.824) | Apparent same cohort; no external validation |
| **Zhu 2025 (7); StageIII** | ROC target not explicit; survival context; CALLY | AUC | 0.604 (0.429–0.809) | Apparent same cohort; no external validation |
| **Liu 2026 [9]; TNBC** | OS; horizon NR; CALLY | AUC | 0.688 (0.603–0.773) | Apparent same cohort; no external validation |
| **Zhu 2025 (7); StageIII** | DFS; Multivariable nomogram containing CALLY | C-index | 0.692 (0.541–0.811) | Author: internal validation; details NR; no external validation |
| **Zhu 2025 (7); StageIII** | DFS 1/3/5/10y; Nomogram | Calibration curves | Qualitative / NR | Apparent same cohort; no external validation |
| **Zhu 2025 (7); StageIII** | DFS 3/5/10y; Nomogram vs CALLY alone | Decision curves | Qualitative / NR | Apparent same cohort; no external validation |
| **Zhu 2025 (7); StageIII** | OS; Multivariable nomogram containing CALLY | C-index | 0.730 (0.586–0.838) | Author: internal validation; details NR; no external validation |
| **Zhu 2025 (7); StageIII** | OS 1/3/5/10y; Nomogram | Calibration curves | Qualitative / NR | Apparent same cohort; no external validation |
| **Zhu 2025 (7); StageIII** | OS 3/5/10y; Nomogram vs CALLY alone | Decision curves | Qualitative / NR | Apparent same cohort; no external validation |
| **Liu 2026 (8); TNBC** | DFS; Multivariable nomogram containing CALLY | C-index | 0.790 (0.619–0.897) | Author: internal validation; details NR; no external validation |
| **Liu 2026 (8); TNBC** | DFS 1/3/5/10y; Nomogram | Calibration curves | Qualitative / NR | Apparent same cohort; no external validation |
| **Liu 2026 (8); TNBC** | DFS 3/5/10y; Nomogram vs CALLY alone | Decision curves | Qualitative / NR | Apparent same cohort; no external validation |
| **Liu 2026 (8); TNBC** | OS; Multivariable nomogram containing CALLY | C-index | 0.808 (0.639–0.909) | Author: internal validation; details NR; no external validation |
| **Liu 2026 (8); TNBC** | OS 1/3/5/10y; Nomogram | Calibration curves | Qualitative / NR | Apparent same cohort; no external validation |
| **Liu 2026 (8); TNBC** | OS 3/5/10y; Nomogram vs CALLY alone | Decision curves | Qualitative / NR | Apparent same cohort; no external validation |
| **Liu 2026 (8); TNBC** | DFS; Nomogram | C-index; abstract conflicting | 0.717 (0.595–0.814) | Apparent same cohort; no external validation |
| **Liu 2026 (8); TNBC** | OS; Nomogram | C-index; abstract conflicting | 0.725 (0.605–0.819) | Apparent same cohort; no external validation |
| **Güney 2026 (3); NACT** | pCR; Delta CALLY alone | AUC | 0.685 (CI NR) | Apparent same cohort; no external validation |
| **Güney 2026 (3); NACT** | pCR; Delta NPAR + delta CALLY score | AUC | 0.755 (0.691–0.815) | Apparent same cohort; no external validation |
| **Güney 2026 (3); NACT** | pCR; Combined score | Optimism-corrected AUC | 0.748 (CI NR) | Bootstrap1000 |
| **Güney 2026 (3); TNBC** | pCR; Combined score | AUC | 0.782 (0.685–0.879) | Apparent same cohort; no external validation |
| **Güney 2026 (3); HER2+** | pCR; Combined score | AUC | 0.764 (0.658–0.870) | Apparent same cohort; no external validation |
| **Güney 2026 (3); HR+/HER2-** | pCR; Combined score | AUC | 0.721 (0.604–0.838) | Apparent same cohort; no external validation |

Performance metrics differ in target, timing, model and validation, so none were pooled. Results and abstract C-index values from Liu are conflicting reports, not independent validations. Brier score measures overall predictive error; a nonsignificant goodness-of-fit test is not proof of calibration. DCA findings are not clinical-impact evidence.

**Table S5. Newcastle–Ottawa assessment: domain and overall scores**

| **Study and analysis** | **S1** | **S2** | **S3** | **S4** | **C1** | **C2** | **O1** | **O2** | **O3** | **Selection** | **Comparability** | **Outcome** | **Total** |
| --- | --- | --- | --- | --- | --- | --- | --- | --- | --- | --- | --- | --- | --- |
| **Karatlı 2025 (1); pCR** | 0 | 1 | 1 | 1 | 1 | 1 | 1 | 1 | 0 | 3/4 | 2/2 | 2/3 | 7/9 |
| **Bahardoust 2026 (5); OS and DFS** | 0 | 1 | 1 | 1 | 1 | 1 | 1 | 1 | 0 | 3/4 | 2/2 | 2/3 | 7/9 |
| **Algın 2026 (4); Stage association** | 1 | 0 | 0 | 1 | 0 | 0 | 2 | 0 | NA | 2/5 | 0/2 | 2/3 | 4/10 |
| **Güneş 2026 (2); Upfront pN positivity** | 1 | 0 | 0 | 2 | 1 | 1 | 2 | 1 | NA | 3/5 | 2/2 | 3/3 | 8/10 |
| **Güneş 2026 (2); Post-NACT ypN positivity** | 1 | 0 | 0 | 2 | 0 | 0 | 2 | 1 | NA | 3/5 | 0/2 | 3/3 | 6/10 |
| **Güneş 2026 (2); Observed nodal burden (both settings)** | 1 | 0 | 0 | 2 | 0 | 0 | 2 | 1 | NA | 3/5 | 0/2 | 3/3 | 6/10 |
| **Zhuang 2024 (6); OS and local-recurrence DFS** | 0 | 1 | 1 | 1 | 1 | 1 | 1 | 1 | 0 | 3/4 | 2/2 | 2/3 | 7/9 |
| **Zhu 2025 (7); OS and DFS** | 0 | 1 | 1 | 1 | 1 | 1 | 1 | 0 | 0 | 3/4 | 2/2 | 1/3 | 6/9 |
| **Liu 2026 (8); OS and DFS** | 0 | 1 | 1 | 1 | 1 | 1 | 1 | 1 | 0 | 3/4 | 2/2 | 2/3 | 7/9 |
| **Güney 2026 (3); Single deltaCALLY vs pCR** | 0 | 1 | 1 | 1 | 0 | 0 | 1 | 1 | 0 | 3/4 | 0/2 | 2/3 | 5/9 |
| **Güney 2026 (3); Combined score vs pCR** | 0 | 1 | 1 | 1 | 1 | 1 | 1 | 1 | 0 | 3/4 | 2/2 | 2/3 | 7/9 |
| **Jimoh 2023 (9); Cancer-only clinicopathology** | 1 | 0 | 0 | 1 | 0 | 0 | 2 | 0 | NA | 2/5 | 0/2 | 2/3 | 4/10 |

Cohort item maxima are 1 each, including separate comparability items C1/C2. Cross-sectional maxima: S1/S2/S3=1 each; S4=2; C1/C2=1 each; O1=2; O2=1; O3 not applicable. Item meanings differ across versions. Initial single-assessor scores only; no independent consensus is claimed. Do not compare raw 9-point and 10-point totals as a common risk scale. Standard cohort NOS and the cited cross-sectional adaptation were used (10, 11)

**Table S6.** GRADE assessment of certainty of evidence for selected key outcomes

| **Outcome** | **Risk of bias** | **Inconsistency** | **Indirectness** | **Imprecision** | **Overall certainty** |
| --- | --- | --- | --- | --- | --- |
| **OS** | Serious | Not serious | Not serious | Serious | **Low** |
| **Combined DFS, exploratory** | Serious | Not serious | Not serious | Serious | **Low** |
| **Baseline CALLY–pCR, HER2-positive cohort** | Serious | Not assessable | Serious | Not seriousᵇ | **Low** |
